# Performance drift is a major barrier to the safe use of machine learning in cardiac surgery

**DOI:** 10.1101/2023.01.21.23284795

**Authors:** Tim Dong, Shubhra Sinha, Ben Zhai, Daniel P Fudulu, Jeremy Chan, Pradeep Narayan, Andy Judge, Massimo Caputo, Arnaldo Dimagli, Umberto Benedetto, Gianni D. Angelini

## Abstract

**Objectives:** The Society of Thoracic Surgeons (STS), and EuroSCORE II (ES II) risk scores, are the most commonly used risk prediction models for adult cardiac surgery post-operative in-hospital mortality. However, they are prone to miscalibration over time, and poor generalisation across datasets and their use remain controversial. It has been suggested that using Machine Learning (ML) techniques, a branch of Artificial intelligence (AI), may improve the accuracy of risk prediction. Despite increased interest, a gap in understanding the effect of dataset drift on the performance of ML over time remains a barrier to its wider use in clinical practice. Dataset drift occurs when a machine learning system underperforms because of a mismatch between the dataset it was developed and the data on which it is deployed. Here we analyse this potential concern in a large United Kingdom (UK) database.

**Methods:** A retrospective analyses of prospectively routinely gathered data on adult patients undergoing cardiac surgery in the UK between 2012-2019. We temporally split the data 70:30 into a training and validation subset. ES II and five ML mortality prediction models were assessed for relationships between and within variable importance drift, performance drift and actual dataset drift using temporal and non-temporal invariant consensus scoring, combining geometric average results of all metrics as the Clinical Effective Metric (CEM).

**Results:** A total of 227,087 adults underwent cardiac surgery during the study period with a mortality rate of 2.76%. There was a strong evidence of decrease in overall performance across all models (p < 0.0001). Xgboost (CEM 0.728 95CI: 0.728-0.729) and Random Forest (CEM 0.727 95CI 0.727-0.728) were the best overall performing models both temporally and non-temporally. ES II perfomed worst across all comparisons. Sharp changes in variable importance and dataset drift between 2017-10 to 2017-12, 2018-06 to 2018-07 and 2018-12 to 2019-02 mirrored effects of performance decrease across models.

**Conclusions:** Combining the metrics covering all four aspects of discrimination, calibration, clinical usefulness and overall accuracy into a single consensus metric improved the efficiency of cognitive decision-making. All models show a decrease in at least 3 of the 5 individual metrics. CEM and variable importance drift detection demonstrate the limitation of logistic regression methods used for cardiac surgery risk prediction and the effects of dataset drift. Future work will be required to determine the interplay between ML and whether ensemble models could take advantage of their respective performance advantages.

**Central message:** ML performance decreases over time due to dataset drift, but remains superior to ES II. Therefore regular assessment and modification of ML models may be preferable.

**Prospective message:** A gap in understanding the effect of dataset drift on the performance of ML models over time presents a major barrier to their clinical application. Xgboost and Random Forest have shown superior performance both temporally and non-temporally against ES II. However, a decrease in model performance of all models due to dataset drift suggests the need for regular drift monitoring.

## Introduction

Recently, the importance of Machine Learning (ML), a branch of Artificial intelligence (AI) has been highlighted as a potential alternative to conventional mortality risk stratification models such as Society of Thoracic Surgeons (STS),[1] and EuroSCORE II (ES II) risk scores,[2] which are prone to miscalibration overtime and poor generalisation across datasets.[1,3] In particular, the ES II, which is based on logistic regression using 18 items of information about the patient, has been shown by numerous studies to display poor discrimination and calibration across datasets with differing characteristics, including but not limited to age,[4] ethnicity[5] and procedures groups.[6–10]

Risk scoring models’ performance are challenged by numerous factors, such as differences in variable definitions, management of incomplete data fields, surgical procedure selection criteria, and temporal changes in the prevalence of patients’ risk factors.[11] ML approaches are increasingly used for prediction in health care research as they have the potential to overcome limitations of linear models. By including pairwise and higher-order interactions and modelling nonlinear effects ML may overcome heterogeneity in procedures and missing data.[1,12] Whilst ML has been shown to be beneficial over conventional scoring systems, the magnitude and clinical influence of such improvements remain uncertain.[2] The ability to counter “performance drift” due to temporal changes in the prevalence of risk factors has also yet to be fully elucidated.

We envisaged that different Machine learning models may perform better for different metrics and that providing a panel of metrics would be important for covering the multifaceted aspects of clinical model performance. The Miller’s law observed that the human working memory is limited to holding on to an average of seven items in the short-term memory.[13] This is particularly relevant in a scenario where a clinician would need to select from a number of ML models based on a panel of performance metrics. The split-attention effect cognition theory indicates that a single integrated source of information enhances knowledge acquisition better than separated sources of information.[14] Therefore, we a consensus approach to metric evaluation by combining the five performance metrics for risk stratification.

We therefore, trained and evaluated 5 supervised ML models to: (1) determine the best ML model in terms of overall accuracy, discrimination, calibration and clinical effectiveness, (2) use variable importance drift as a measure for detecting dataset drift and (3) verify suspected dataset drift by assessing the relationship between and within performance drift, variable importance drift and dataset drift (e.g. due to changing case-mix[15]) across ML and ES II approaches.[16]

## Methods

### Dataset and Patient Population

The study was performed using the National Adult Cardiac Surgery Audit (NACSA) dataset, which comprises data prospectively collected by National Institute for Cardiovascular Outcome Research on all cardiac procedures performed in all NHS hospitals and some private hospitals across the UK.[17] Patients undergoing cardiac surgery between 1 Jan 2012 and 31 Mar 2019 were included. Missing and erroneously inputted data in the dataset were cleaned according to the National Adult Cardiac Surgery Audit Registry Data Pre-processing recommendations.[18] Generally, for any variable data that were missing, it was assumed that the variable was at baseline level, i.e., no risk factor was present. Missing patient age at the time of surgery was imputed as the median patient age for the corresponding year. Data standardization was performed by subtracting the variable mean and dividing by the standard deviation values.[19]

The dataset was split into two cohorts: Training/Validation (n = 157196; 2012-2016; Table S1) and Holdout (n = 69891; 2017-2019; Table S2). The primary outcome of this study was in-hospital mortality.

The study was part of a research project approved by the Health Research Authority (HRA) and Health and Care Research Wales. As the study included retrospective interrogation of the NICOR database, the need for individual patient consent was waived (HCRW) (IRAS ID: 278171) in accordance with the research guidance. The study was performed in accordance with the ethical standards as laid down in the 1964 Declaration of Helsinki and its later amendments.

### Baseline Statistical analysis

Continuous variables are compared using non-parametric Wilcoxon rank-sum tests, whilst categorical variables are compared using Pearson’s χ ^2^tests or Fisher’s exact tests as appropriate.

Scikit-learn v0.23.1 and Keras v2.4.0 were used to develop the models and to evaluate their discrimination, calibration and clinical effectiveness capabilities. Statistical analyses are conducted using STATA-MP version 17 and R v4.0.2.[20] Anova Assumptions were checked using R rstatix package.

### Model Development

In our study, we trained five supervised ML risk models based on the ES II preoperative variable set (Table S3). Those five models included Logistic Regression, Neural Network (Neuronetwork / NN),[19] Random Forest (RF),[21] Weighted Support Vector Machine (SVM),[22] and Xgboost[23].[17] ES II score was calculated for baseline comparison. Internal validation was performed using fivefold cross-validation on the Training/Validation dataset (2012-2016). External validation was performed on the Holdout dataset (2017-2019).[16] Each model calculated the probability of surgical mortality for each patient. One thousand bootstrap samples were taken for all metrics. Further details on model development can be found in Supplementary Materials, section: Model Specification.

### Assessment of model performance

The Area Under the Curve (AUC) performances of all variant models were evaluated, and the ROC curves plotted.[24] As a sensitivity analysis, we excluded the True Negative Rate from the performance evaluation, by calculating the F_*1*_ score.[25] This metric adjusts for the biased effect due to high proportion of alive outcome samples. Decision Curve net benefit index is used to test clinical benefit.[26] 1 - Expected Calibration Error (ECE) was used to determine calibration performance, with higher values being better.[27] The adjusted Brier score (1 – Brier) was used without the normalization term,[28] but with higher values indicating better discrimination and calibration performance.

To determine the best model in terms of both discrimination and calibration, we took the geometric average of AUC, F1,[25] Decision Curve net benefit (Treated + Untreated), 1 – ECE and 1 – Brier. Geometric average has previously been found to be effective for summarising metrics for temporal based model calibration[29]. This metric is robust to outliers,[30] and is preferable for aggregation compared to the weighted geometric mean.[31] The arithmetic average was used for Decision Curve net benefit over all thresholds as a measure of overall net benefit, before geometric averaging, since values can be negative. We proposed a new metric using the combined geometric average results of all metrics, named Clinical Effective Metric (CEM). An overview of the model and evaluation design is shown in Figure 1.

**Figure 1.**
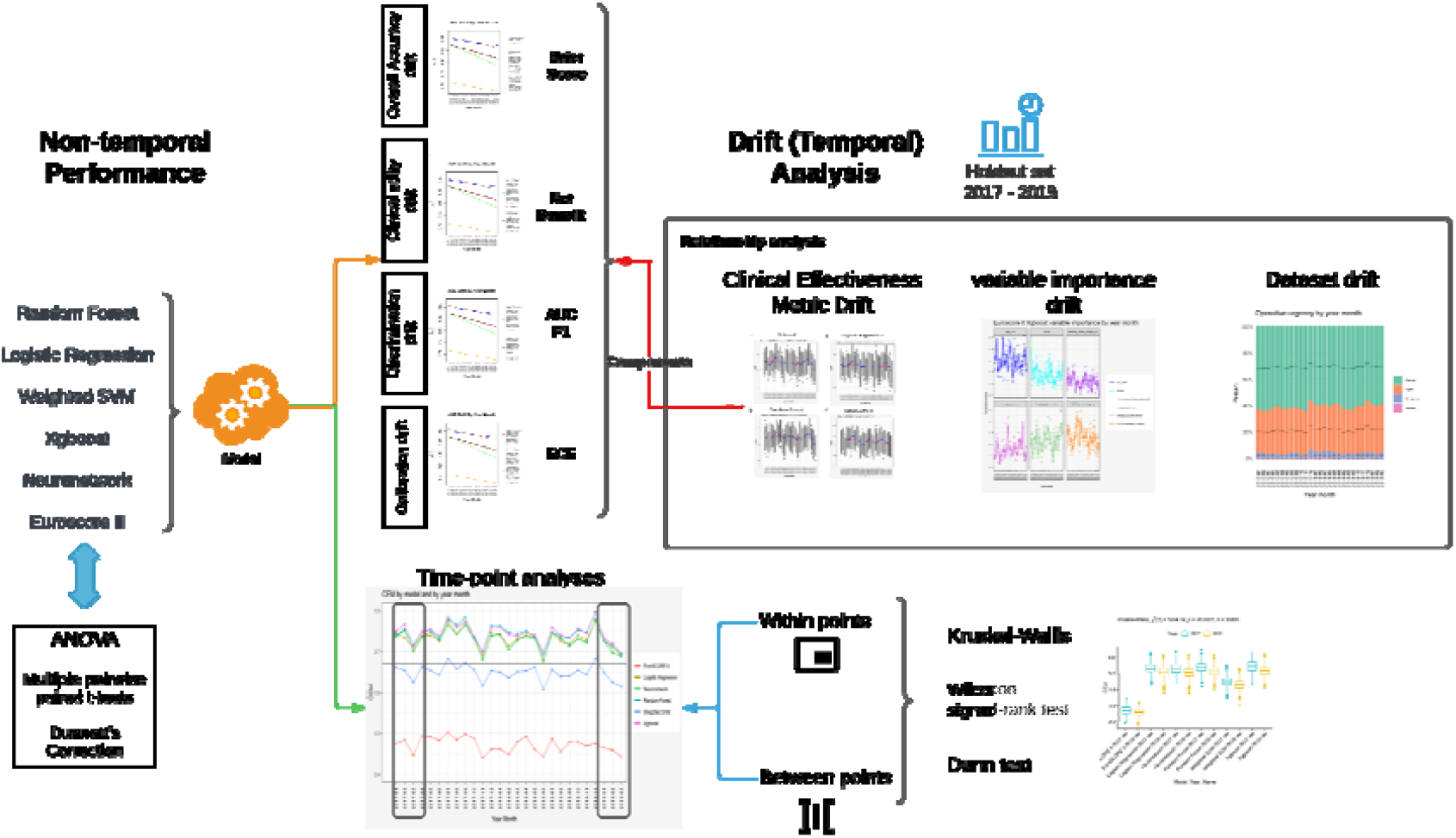
Design overview of the study; non-temporal performance and drift (temporal) analyses are performed; drift in discrimination, calibration, clinical utility, dataset and variable importance are assessed; time point assessments are performed for CEM; drifts in component metrics of CEM are evaluated.

### Baseline non-temporal performance

Non-temporal comparison of models was conducted as a baseline, using all data across the Holdout period. Differences across models were tested using repeated measures One-Way Anova and Bonferroni Corrected multiple pairwise paired t-tests; this was followed by Dunnett’s Correction for multiple comparisons, with the best overall performing model as control. ANOVA assumptions for outliers were checked. Normality assumptions were checked using the Shapiro-Wilk test.[32]

### Drift Analysis

#### CEM Regression trends

Geometric CEM mean of 1000 bootstraps for each model against month of the year was calculated as well as 95% CI and the results were plotted to compare trends across models. The models were compared by fitting multiple linear regression lines across year months for CEM.

To check for normality assumptions, we plotted the histogram and a QQ plot of residuals before applying linear regression.[33] We also checked for homogeneity of residual variance (homoscedasticity) by plotting a scale-location plot i.e. the square root of standardised residual points against values of the fitted outcome variable.[34] For model metrics that do not satisfy these assumptions, the Seasonall Kendall Test (non-parametric) was used instead.

#### Analysis within first 3 months of 2017 and 2019

Differences in CEM across models at two time-points were independently tested using Kruskal-Wallis Test and Bonferroni Corrected paired samples Wilcoxon test (Wilcoxon signed-rank test). The two time-points were the first three months of 2017 and 2019, respectively. This was followed by the Dunn test for non-parametric multiple comparisons of models at each of the two-time points, with the best overall performing model as a baseline. ANOVA assumptions for outliers were checked. Normality assumptions were checked using the Shapiro-Wilk test.[32]

#### Analysis between first 3 months of 2017 and 2019

Differences in models’ CEM across the first three months of 2017 and 2019 were tested using Kruskal-Wallis Test and paired samples Wilcoxon test (Wilcoxon signed-rank test). Dunn test was used to determine the magnitude and evidence of change across the two-time points for each model. ANOVA assumptions for outliers were checked. Normality assumptions were checked using Kolmogorov-Smirnov Test.

#### Analysis of discrimination, calibration, clinical utility and overall accuracy drift

As a sensitivity analysis, we analysed performance drift in terms of component metrics within CEM. Discrimination (AUC), positive outcome discrimination (F1 score), calibration (1 - ECE), clinical utility (net benefit), Adjusted Brier score (overall accuracy of prediction probability) were assessed by fitting multiple (model) linear regression lines across year month for each metric, respectively.

To check for normality assumptions, we plotted the histogram and a QQ plot of residuals before applying linear regression.[33] We also checked for homogeneity of residual variance (homoscedasticity) by plotting a scale-location plot i.e. the square root of standardised residual points against values of the fitted outcome variable.[34] For model metrics that do not satisfy these assumptions, the Seasonall Kendall Test (non-parametric) is used instead.

#### Analysis of variable importance drift

Variable importance drift was assessed for the best performing model. For each year month of the Holdout dataset, 5-fold nested cross-validation was performed to derive importance of each ES II variable in the model’s decision making. The geometric mean of 5-fold importance at each time point was plotted along with the importance of each of the 5 folds. The SHAP mean absolute magnitude of importance was used.[35,36] Loess smoothing was used to simplify the visual representation. Line plots of the top six important variables were used as sensitivity analysis.

#### Dataset drift

Dataset drift across year month was visualised using a stacked bar plot for the top three variables as identified by SHAP variable importance. Continuous variables were binned into intervals to enable ease of analysis.

## Results

### Baseline patient characteristics

A total of 227,087 procedures of adults from 42 hospitals were included in this analysis. This followed the removal of 3,930 congenital cases, 1,586 transplant and mechanical support device insertion cases and 3,395 procedures missing information on mortality (Table 1). There were 6,258 deaths during the study period (mortality rate of 2.76%).

**Table 1.** Patient Demographics. Summary of cleaned Euroscore II Variables. Variables are for the time period 2012 – 2019. Records with missing mortality status were excluded.

| Variable | Mortality Status |  | p-value<sup>2</sup> |
| --- | --- | --- | --- |
|  | 0, N = 220,829<sup>1</sup> | 1, N = 6,258<sup>1</sup> |  |
| **Age (years), mean (SD)** | 67.53 (11.23) | 70.77 (11.42) | <0.001 |
|  | 0, N = 220,829<sup>1</sup> | 1, N = 6,258<sup>1</sup> |  |
| **NYHA, n (%)** |  |  | <0.001 |
| 0 – I | 48,625 (22%) | 1,055 (17%) |  |
| 1 – II | 96,888 (44%) | 1,609 (26%) |  |
| 2 – III | 64,049 (29%) | 2,228 (36%) |  |
| 3 – IV | 11,267 (5.1%) | 1,366 (22%) |  |
| **Renal impairment, n (%)** |  |  | <0.001 |
| 0 - Normal | 103,196 (47%) | 1,704 (27%) |  |
| 1 - Moderate | 92,411 (42%) | 2,451 (39%) |  |
| 2 - On Dialysis | 2,187 (1.0%) | 330 (5.3%) |  |
| 3 - Severe | 23,035 (10%) | 1,773 (28%) |  |
| **Chronic lung disease, n (%)** | 26,644 (12%) | 1,211 (19%) | <0.001 |
| **Poor mobility, n (%)** | 8,305 (3.8%) | 514 (8.2%) | <0.001 |
| **Previous cardiac surgery, n (%)** | 12,012 (5.4%) | 1,141 (18%) | <0.001 |
| **LV function** |  |  | <0.001 |
| 0 - Good (>50%) | 184,721 (84%) | 4,706 (75%) |  |
| 1 - Moderate (31-50%) | 30,608 (14%) | 1,089 (17%) |  |
| 2 - Poor (21-30%) | 4,241 (1.9%) | 318 (5.1%) |  |
| 3 - Very Poor (≤20%) | 1,259 (0.6%) | 145 (2.3%) |  |
| **Pulmonary hypertension, n (%)** |  |  | <0.001 |
| 0 – PA Systolic (<31mmHg) | 201,643 (91%) | 5,000 (80%) |  |
| 1 – PA Systolic (31-55 mmHg) | 13,126 (5.9%) | 705 (11%) |  |
| 2 – PA Systolic (>55mmHg) | 6,060 (2.7%) | 553 (8.8%) |  |
| **CCS class 4 angina, n (%)** | 18,370 (8.3%) | 956 (15%) | <0.001 |
| **Urgency, n (%)** |  |  | <0.001 |
| 0 - Elective | 141,617 (64%) | 2,442 (39%) |  |
| 1 - Urgent | 72,090 (33%) | 2,134 (34%) |  |
| 2 - Emergency | 6,533 (3.0%) | 1,230 (20%) |  |
| 3 - Salvage | 589 (0.3%) | 452 (7.2%) |  |
| **Weight of the intervention, n (%)** |  |  | <0.001 |
| 0 – Isolated CABG | 111,243 (50%) | 1,546 (25%) |  |
|  | 0, N = 220,829<sup>1</sup> | 1, N = 6,258<sup>1</sup> |  |
| 1 – Single non-CABG | 62,568 (28%) | 2,153 (34%) |  |
| 2 – 2 Procedures | 42,649 (19%) | 2,108 (34%) |  |
| 3 – 3 Procedures | 4,369 (2.0%) | 451 (7.2%) |  |
| **Diabetes on insulin, n (%)** | 12,818 (5.8%) | 453 (7.2%) | <0.001 |
| **Female gender, n (%)** | 59,467 (27%) | 2,328 (37%) | <0.001 |
| **Recent myocardial infarct, n (%)** | 43,316 (20%) | 1,594 (25%) | <0.001 |
| **Critical preoperative state, n (%)** | 7,255 (3.3%) | 1,382 (22%) | <0.001 |
| **Extracardiac arteriopathy, n (%)** | 22,327 (10%) | 1,215 (19%) | <0.001 |
| **Active endocarditis, n (%)** | 5,816 (2.6%) | 493 (7.9%) | <0.001 |
| **Surgery on thoracic aorta, n (%)** | 9,070 (4.1%) | 896 (14%) | <0.001 |
| **Euroscore II, mean (SD)** | 0.03 (0.04) | 0.12 (0.14) | <0.001 |

<sup>1</sup>Mean (SD) or Frequency (%)
<sup>2</sup>Wilcoxon rank sum test; Pearson's Chi-squared test

### Baseline non-temporal performance

No extreme outliers were found. The CEM scores were normally distributed for all three models except Xgboost, as assessed by Shapiro-Wilk’s test (p > 0.05). A histogram plot of the Xgboost CEM values did not show substantial deviation from the normal distribution. There was strong evidence of a difference across all models p < 0.0001 (Table S4 and Figure S1). Table 2 shows that Xgboost (CEM 0.728 95CI (95% confidence interval): 0.728-0.729) and RF (CEM 0.727 95CI 0.727-0.728) are the best overall performing models, with moderate to strong evidence (non-overlapping CI) of the former outperforming the latter. This was followed by LR, NN, SVM then ES II. Dunnett’s test showed that there was moderate to strong evidence that Xgboost was superior to all other models (p < 0.001) (Table 3). Xgboost performance was least different from RF, but most different from ES II (CEM difference 0.0009 vs. 0.1876).

**Table 2.** Geometric Mean of Individual metrics for each model in the Holdout set; CEM refers to Clinical Effective Metric; Standard deviation and 95% CI are shown for CEM. 1000 bootstrap samples were used to derive geometric mean of each metric; adjusted 1 - ECE and 1 - Brier score values are shown; net benefit is average absolute overall benefit across all thresholds.

| Model Category | 1 - ECE | AUC | 1 - Brier | F1 | Net Benefit | CEM.Mean | CEM.sd | CEM.n | CEM lower CI | CEM upper CI |
| --- | --- | --- | --- | --- | --- | --- | --- | --- | --- | --- |
| EuroSCORE II | 0.641 | 0.800 | 0.814 | 0.240 | 0.461 | 0.541 | 0.004 | 1000 | 0.540 | 0.541 |
| LR | 0.997 | 0.819 | 0.976 | 0.264 | 0.902 | 0.717 | 0.005 | 1000 | 0.717 | 0.717 |
| NN | 0.997 | 0.813 | 0.976 | 0.259 | 0.901 | 0.713 | 0.006 | 1000 | 0.713 | 0.714 |
| RF | 0.996 | 0.835 | 0.976 | 0.277 | 0.904 | 0.727 | 0.005 | 1000 | 0.727 | 0.728 |
| Weighted SVM | 0.775 | 0.819 | 0.916 | 0.257 | 0.685 | 0.634 | 0.005 | 1000 | 0.634 | 0.634 |
| Xgboost | 0.996 | 0.834 | 0.976 | 0.279 | 0.904 | 0.728 | 0.005 | 1000 | 0.728 | 0.729 |

**Table 3.**
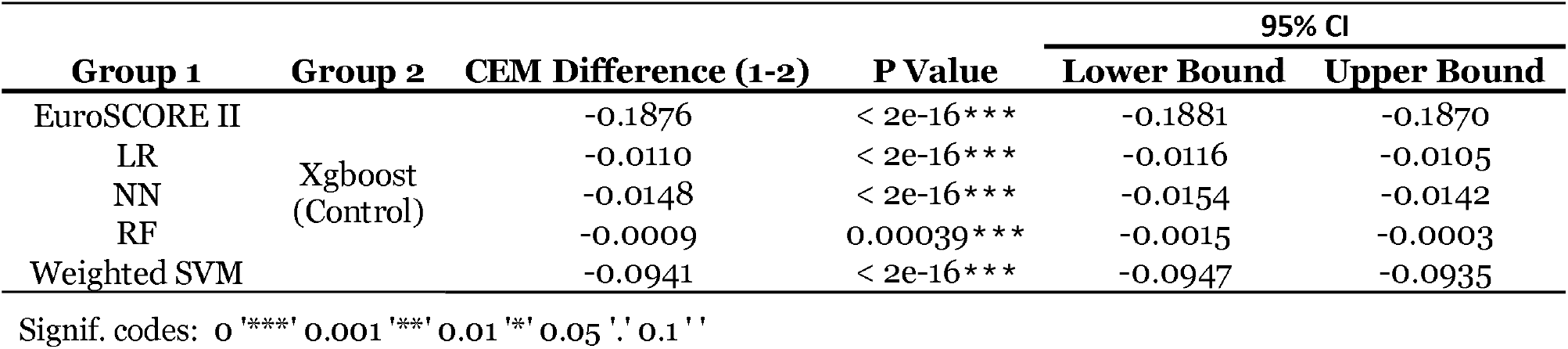
Dunnett’s test with Xgboost as control and the rest of the models as comparison; 95% family-wise confidence level are shown as well as mean difference in CEM and p-values.

Sensitivity analysis of CEM component metrics shows that the adjusted Brier score was unable to distinguish Xgboost, RF, NN and LR (Table 2, all 0.976). AUC performance was best for Xgboost (0.834) and RF (0.835). F1 score showed that Xgboost performed best followed by RF (0.279 vs. 0.277). LR and NN (adjusted ECE: both 0.997) showed better calibration performance than RF and Xgboost (adjusted ECE: both 0.996). Net Benefit overall was best for Xgboost and RF (both 0.904).

### Drift Analysis Overall CEM

Figure 2a shows that Xgboost and RF were candidates for the best overall CEM performance across year month. There was minor evidence of LR outperforming NN across time. Seasonal fluctuations were observed. ES II performed worst across time followed by SVM.

**Figure 2.**
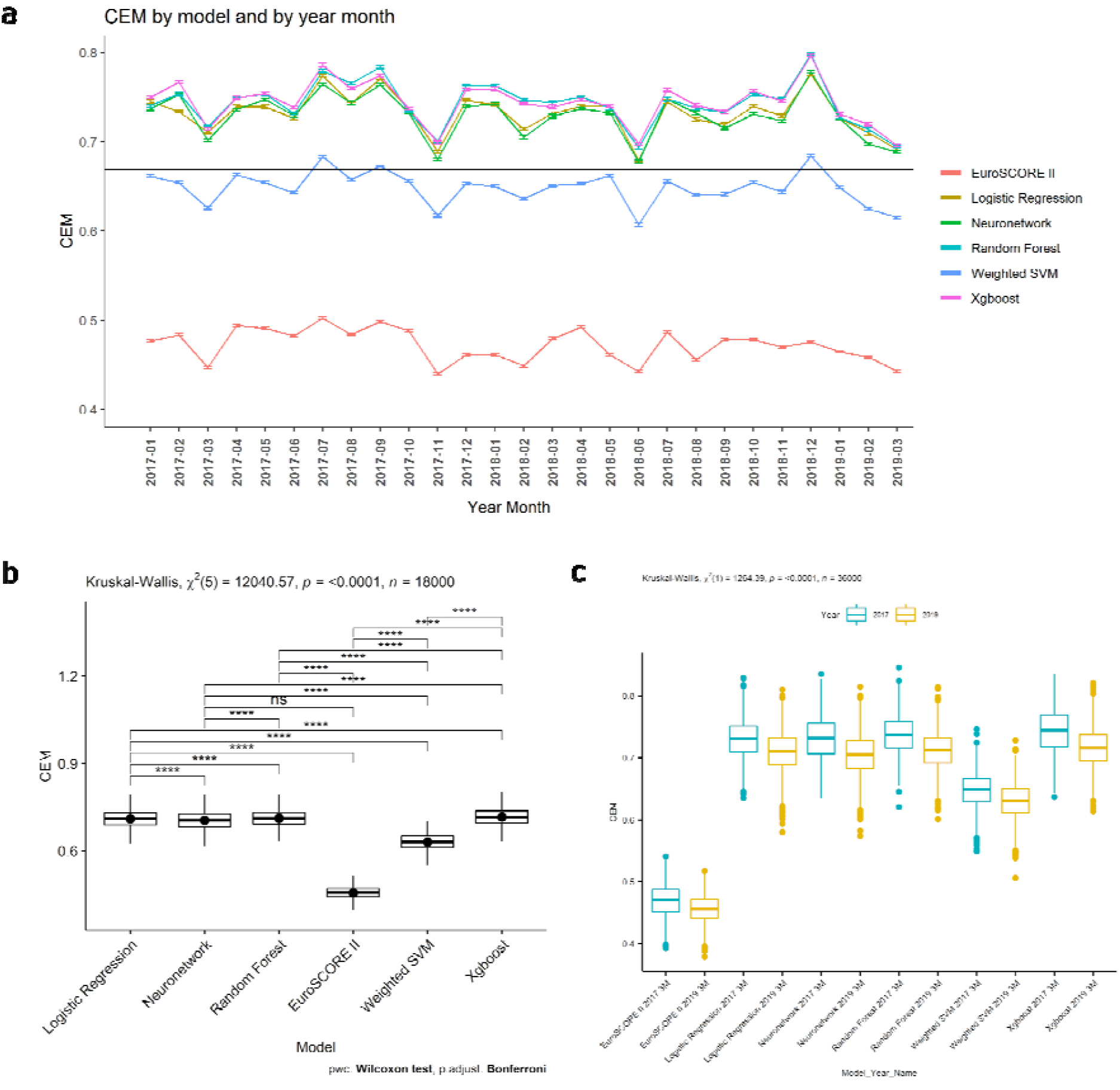
a) Plot of CEM by model and by year month; geometric mean of 1000 bootstraps at each time point is shown as is 95% CI; horizontal line represents the CEM geometric mean of all models; b) Box plot of difference in models’ CEM across first three months of 2017 and 2019; Kruskal-Wallis results for CEM across the time points are shown; c) Paired samples Wilcoxon test (Wilcoxon signed-rank test) for first 3 months of 2019 bootstrap CEM values; p-values are adjusted using the bonferroni method.

There was strong evidence of a decrease in overall performance across all models (p < 0.0001). Linear regression plots showed that Xgboost had the best starting CEM (intercept = 0.755 vs. 0.753 (RF), 0.742 (LR), 0.741 (NN)), but rate of performance decrease (slope: - 0.000720) was less than NN (slope: -0.00083) and greater than RF (-0.000685) and LR (-0.000696; Figure 3a-c and Figure S2.1).

**Figure 3.**
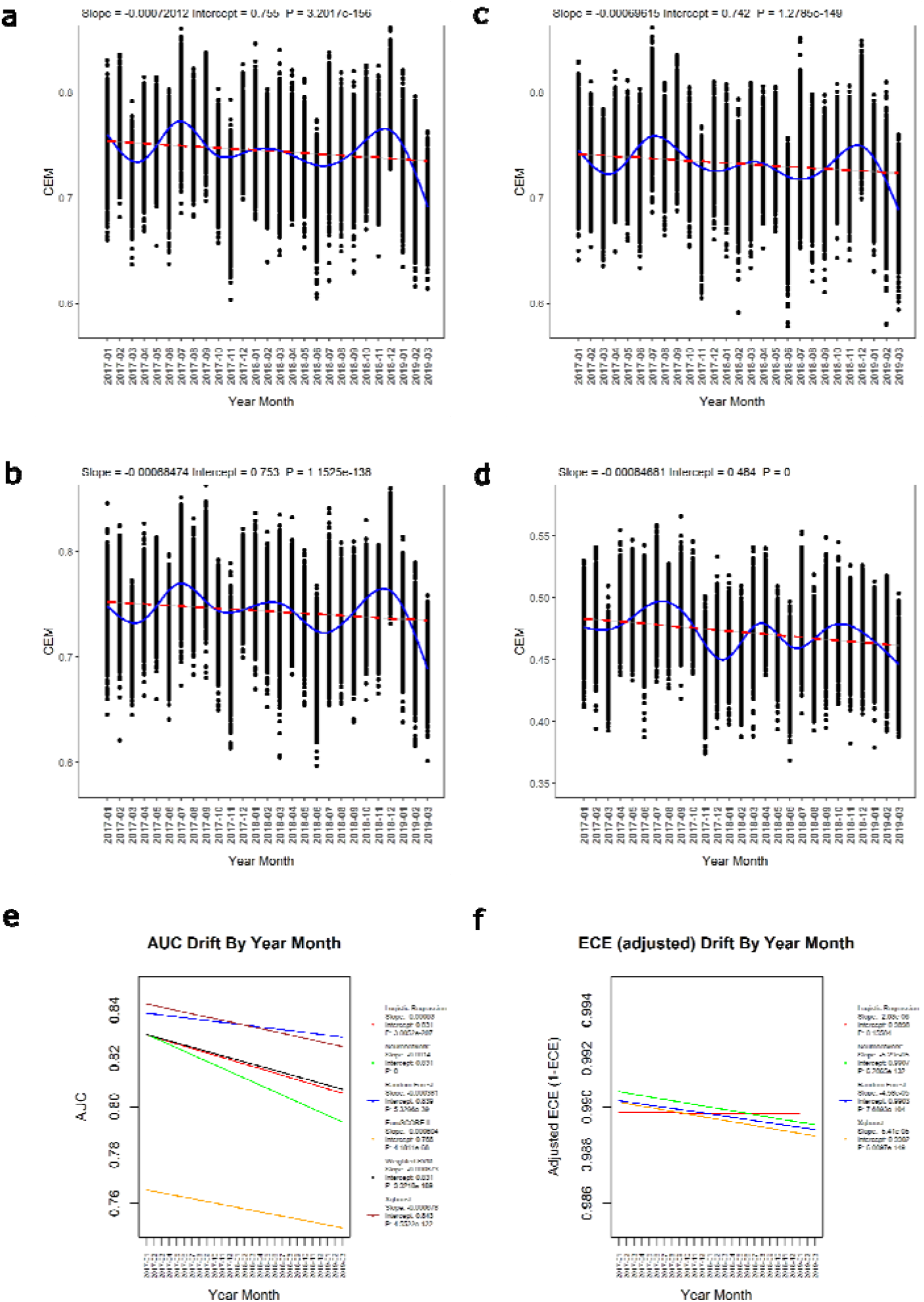
Plot of CEM by model (a. Xgboost; b. Random Forest; c. Logistic Regression; d. EuroSCORE II) and by year month; geometric mean of 1000 bootstraps at each time point is shown; red dotted line shows linear regression; blue line shows Generalised Additive Model fit (GAM); parameters and p-value for linear regression are shown; e) Discrimination (AUC) performance drift by year month; linear regression lines are plotted for each model with slope, intercept and p-values displayed in legend; f) Calibration (adjusted ECE) performance drift by year month; linear regression lines are plotted for each model with slope, intercept and p-values displayed in legend; SVM and ES II are removed to enable clearer separation of models with similar performance.

By March 2019, the overall CEM performance ranking was not changed, with Xgboost performing best, followed by RF, LR and then NN. ES II (intercept: 0.484, slope:-0.000847) performed worst in terms of starting CEM and rate of performance decrease, followed by SVM (intercept:0.658; slope: -0.000625; Figure 3d and S2.2). Normality and homogeneity assumptions were satisfied for all model CEM values as checked by QQ plot of residuals and scale-location plot (Supplementary Materials, Figure S2.3).

### Analysis within first 3 months of 2017

No extreme outliers were found for models’ CEM values in the first three month of 2017. The CEM scores were non-normally distributed for all models(p < 0.05). There was strong evidence of a difference across all models (p < 0.0001; Table 2b and Figure S3). Dunn test showed strong evidence of Xgboost having the best overall performance (Table S6, p < 0.0001), followed by RF, NN and then LR (CEM difference to Xgboost: -0.0076, -0.0124 and - 0.0138, p < 0.0001). EuroSCORE II performed worst followed by Weighted SVM (CEM difference to Xgboost: -0.2739, -0.0961, p < 0.0001).

### Analysis within the first 3 months of 2019

No extreme outliers were found for models’ CEM values in the first three month of 2019. The CEM scores were non-normally distributed for 50% of models(p < 0.05). There was strong evidence of a difference across all models (p < 0.0001; Table S7 and Figure 2b). Dunn test showed strong evidence of Xgboost having the best overall performance (Table S8, p < 0.05), followed by RF, LR and then NN (CEM difference to Xgboost: -0.0032, -0.0055 and -0.0108, p < 0.05). EuroSCORE II performed worst followed by Weighted SVM (CEM difference to Xgboost: -0.2594, -0.0856, p < 0.0001).

### Analysis between first 3 months of 2017 and 2019

No extreme outliers were found for models’ CEM values in the first three months of 2017 and 2019. The CEM scores were non-normally distributed for the first 3 months of 2017 and 2019, as assessed by Kolmogorov-Smirnov Test (p < 0.05). There was strong evidence of an overall difference across the two-time points (p < 0.0001; Table S9 and Figure S4). There was strong evidence of a difference across the two-time points for each individual model (p < 0.05; Figure 2c and Table S10). Xgboost retained the best overall performance across the time points examined. This model showed the largest decrease in CEM performance (Median difference: 0.0288, p < 0.0001), followed by NN, RF and then LR (Median difference: 0.0272, 0.0244, 0.0205, p < 0.0001). Following a performance decrease from 2017 to 2019, Xgboost still had the best overall performance with RF being second best (Median CEM 0.716, 0.713) Although NN had better starting performance than LR, the larger performance drift resulted in NN having a lower overall performance than LR at 2019 (0.705 vs. 0.710). However, although performance drift was smaller, LR’s CEM performance never exceeded RF (0.710 vs. 0.713). EuroSCORE II showed the least performance drift followed by Weighted SVM (Median difference: 0.0142, 0.0183, p < 0.05), but both performed worst in terms of absolute CEM.

### Analysis of discrimination, calibration and clinical effectiveness drift

#### Discrimination

##### AUC

Linear regression plots show that Xgboost has the best starting AUC (intercept = 0.843 vs. 0.839 (RF), 0.831 (LR, NN, SVM)), but rate of performance decrease was greater than RF and ES II (slope: -0.000678 vs. -0.000381, -0.000604; Figure 3e). By March 2019, Xgboost AUC had decreased below RF, resulting in RF being the best performing model, followed by Xgboost, SVM, LR and then NN. NN showed the largest rate of AUC decrease followed by LR and SVM (slope: -0.0014, -0.00093, -0.000873). ES II performed worst in terms of AUC across all time points (intercept: 0.766). There was a strong evidence of decrease in AUC performance across all models (p < 0.0001). Normality and homogeneity assumptionswere satisfied for all model AUC values as checked by QQ plot of residuals and scale-location plot (Figure S5).

##### F1 score

The best performing model across all Holdout time periods was Xgboost, followed by RF, LR, NN, SVM and then ES II. There was strong evidence of a decrease in F1 performance across all models (p < 0.0001). For more details, see Supplementary Materials, section: Positive outcome discrimination.

##### Calibration

Linear regression plots showed that NN has the best starting adjusted ECE (intercept = 0.9907 vs. 0.9903 (RF), 0.9902 (Xgboost), 0.9898 (LR)) but rate of performance decrease was greater than LR and RF (slope: -5.29e-5 vs. -2.93e-6, -4.58e-5; Figure 3f). By March 2019, NN adjusted ECE had decreased below LR, resulting in LR being the best performing model, followed by NN, RF and then Xgboost. While SVM and ES II had lower rates of adjusted ECE decrease (slope: -0.000251, -0.000479), the calibration performance was much lower at all time points compared to the other models (Figure S6). There was strong evidence of decrease in adjusted ECE performance across all models (p < 0.0001), except LR (p > 0.05). Normality and homogeneity assumptions were satisfied for all model adjusted ECE values as checked by QQ plot of residuals and scale-location plot (Figure S7).

#### Clinical Effectiveness

Linear regression plots showed that Xgboost has the best starting net benefit (intercept = 0.9051 vs. 0.9043 (RF), 0.9035 (NN, LR)) but rate of performance decrease was greater than RF (slope: -5.68e-5 vs. -2.5e-6; Figure 4a), but slower than LR (-9.38e-5) and even slower than NN (-0.000145).

**Figure 4.**
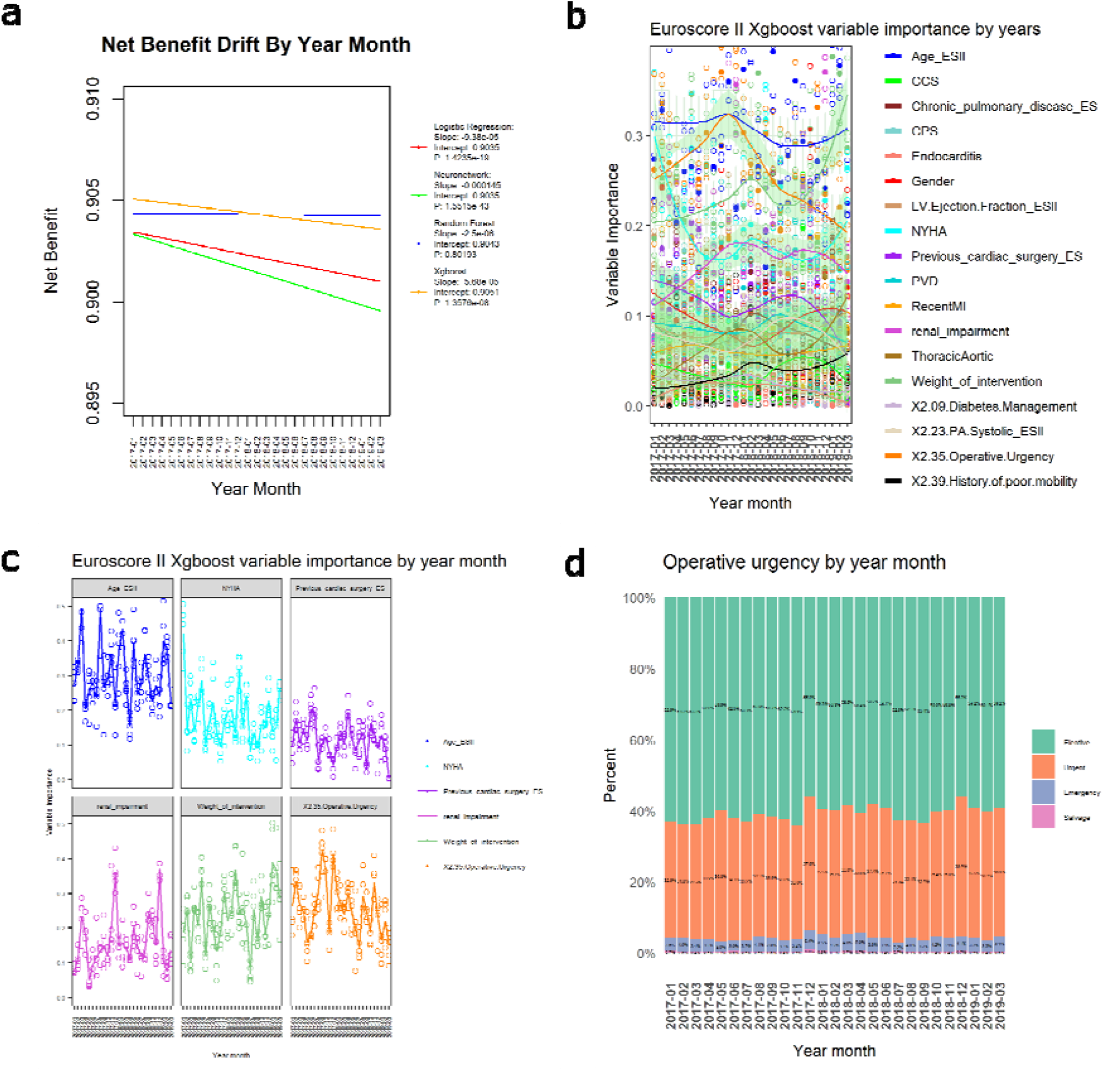
a) Clinical effectiveness (net benefit) performance drift by year month; linear regression lines are plotted for each model with slope, intercept and p-values displayed in legend; SVM and ES II are removed to enable clearer separation of models with similar performance; b) SHAP variable importance drift for 27 month of Holdout set; solid dots show geometric mean values of 5 fold cross validation; smoothed loess lines are plotted, with green bands showing 95% confidence intervals; c) SHAP variable importance drift for 27 month of Holdout set for top six most important variables; trends are unsmoothed; d) Operative urgency dataset drift across year month for Holdout set; percentages of each category are shown for each time point.

By March 2019, Xgboost net benefit had decreased below RF, resulting in RF being the best performing model, followed by Xgboost, LR and NN. ES II showed the largest rate of net benefit decrease and performed worst across all time points followed by SVM (intercept: 0.314, 0.690; slope: -0.000846, -0.000364; Figure S8). There was strong evidence of a decrease in net benefit performance across all models (p < 0.0001), except RF (p > 0.05). Normality and homogeneity assumptions were satisfied for all model net benefit values as checked by QQ plot of residuals and scale-location plot (Figure S9).

#### Accuracy of prediction probability

By March 2019, Xgboost was the best model followed by RF, LR and then NN. ES II performed worst in terms of Adjusted Brier and rate of decrease, followed by SVM. There was strong evidence of a decrease in Adjusted Brier performance across all models (p < 0.0001), except Xgboost and RF. For more details, see Supplementary Materials, section: Accuracy of prediction probability.

#### Analysis of variable importance drift

SHAP mean absolute magnitude of importance was used to measure variable importance drift for the best temporal and non-temporal model (Xgboost). Smoothed trend lines showed substantial drift in numerous variables, including the most important variables: age, operative urgency, the weight of intervention, NYHA, renal impairment and previous cardiac surgery (Figure 4b). Sensitivity analysis showed a substantial drift in variable importance across the Holdout set for all six variables (Figure 4c). When compared with CEM performance drop between 2017-10 to 2017-12 and between 2018-06 to 2018-07 (Figure 3 GAM line), it could be seen that the CEM decrease was mirrored by decreases in the importance of the top variables: age and operative urgency at these time periods (Figure 4c). A decrease in CEM performance in the three months of 2019 was likely to be at least partly contributed to by the sudden rise in importance of the weight of intervention (Figure 3 and Figure 4b, 4c).

#### Dataset drift across time

Dataset drift is observed throughout the Holdout time periods for operative urgency with sharp drifts observed across all categories between 2017-11 (YYYY-MM) to 2017-12 and between 2018-06 and 2018-07 (Figure 4d). Dataset drift was observed across the Holdout time periods for patient age groups above and below 60 (Figure S15), with marked data drifts observed between 2017-10 to 2017-11 and between 2018-07 to 2018-08. Dataset drift was observed across the Holdout time periods for Weight of intervention (Figure S16). Sharp dataset drifts were observed for the Single non-CABG and 3 procedures category between 2018-12 to 2019-02.

## DISCUSSION

The main finding of the study was that Xgboost performed best followed by RF, LR and then NN when all metrics are simultaneously considered, both temporally and non-temporally. Furthermore, EuroSCORE II substantially underperformed against all ML models across all comparisons and presents an urgent need to replace this score. By first combining all metrics and then analysing the temporal drift of each metric individually, we were able to determine the contribution of individual metrics to the overall performance drift of each model. We found strong evidence that all models showed a decrease in at least 3 of the 5 individual metrics within CEM. This demonstrated the importance for clinicians and ML governance teams to actively monitor the effects of dataset drift (as explained later) on “Big Data” models that are prepared for or being clinically used in order to minimise the risk of harm to patients.

“Big data” refers to large and detailed datasets that are suited to ML analyses rather than traditional statistical analyses.[37,38] This is increasingly utilised in healthcare. These analyses can inform, personalise and potentially improve care.[37,39,40] Despite growing interest[41] in ML and healthcare data linkage initiatives such the Health Informatics Collaborative (HIC),[42] there have been limited reports of usage within cardiac surgery,[43– 45] with one of the main reasons being a lack of understanding by clinicians of the underlining processes.[46]

As more countries follow in the steps of the U.S. to deploy ML to the medical settings,[47] it becomes increasingly critical that clinicians and ML governance teams are adequately prepared for situations in which ML systems fail to perform their intended functions.[48] A major factor in ML malfunction is “Dataset Drift”, where ML performance declines due to a mismatch between the data on which the model was trained and the new unseen data to which the model is applied.[49] Several factors have been reported to influence dataset drift, including changes in technology, demographics, and patient or clinician behaviour.[48]

In our previous systematic review, we found that despite ML models achieving better discriminatory ability than traditional LR approaches, few cardiac surgery studies assessed calibration, clinical utility, discrimination and dataset drift collectively; these aspects should be assessed to determine the clinical implications of ML.[2] While calibration drift over time is well documented amongst EuroSCORE and logistic regression models for hospital mortality, the susceptibility of competing ML modelling methods to dataset drift has not been well studied in cardiac surgery.[50]

This study heeds to the call for additional metrics to address the lack of sensitivity of the most commonly used C-statistic and calibration slope in capturing the advantage of ML models,[51] by demonstrating the use of a consensus score[19,52–55] named CEM to take into account numerous metrics that have been found to be beneficial, covering overall accuracy,[51] discrimination, calibration and clinical utility. This study showed invariance in model ranking for the CEM in both temporal and non-temporal analyses, indicating there is value for this consensus scoring approach in performance drift evaluation.

The current study also addresses the gap in understanding the effect of dataset drift on the performance of ML and traditional models over time, which presents a barrier to their clinical application. The shift in best performing AUC and net benefit model between Xgboost and RF, and between NN and LR for “adjusted ECE” demonstrates that comparison of models at a single time point was insufficient to understand the clinical limitations of ML models and at least two-time points should be considered.

Our study has also found that although RF shows comparable discrimination (AUC) and clinical utility (net benefit) performance across time, the reason for Xgboost’s superior overall temporal performance was in its better overall accuracy (Adjusted Brier) and positive outcome discrimination (F1). F1 score is often overlooked, but is especially important in cardiac surgery datasets, whereby the incidence for the outcome of interest is typically very low and introduces bias in the performance evaluation, when AUC is used. We found that RF performed second best overall. Unlike Xgboost, RF performed better in terms of resistance to drift in AUC and net benefit, suggesting that further work is required to determine whether the synergistic (ensemble) effects across models are beneficial for improving cardiac surgery risk prediction. Although Xgboost is currently the best temporal and non-temporal model for the National Adult Cardiac Surgery Audit dataset, periodic monitoring of performance drift for each yearly revision of this dataset should be mandated to determine whether or not performance is overtaken by RF, and if so, at what point in time this happens.[48] As all models showed strong evidence of decrease in overall performance from 2017-01 to 2019-03, further work will be required to develop either better performing models or models that are less susceptible to performance drift.

We have demonstrated that by associating relationships between smoothed[56] and unsmoothed trend lines for CEM performance and ES II variable importance, that it was possible to detect subtle dataset drifts that could result in model performance drifts. Our findings of variable importance and dataset drift between 2017-10 to 2017-12, between 2018-06 to 2018-07 and between 2018-12 to 2019-02 are likely to reflect seasonality changes and mirrored effects of sharp drifts in CEM performance across models. The detection of dataset drift was verified by checking for actual drifts in the dataset variables. A non-cardiac surgery study has used actual dataset drift to check for variable importance detected dataset drift.[50] However, drift in the actual dataset was only analysed across two data points,[50] without consideration for smoothed and unsmoothed relationships across performance, variable importance and actual variable incidence. The current study provides the foundations for which further work analysing ML performance drift are recommended to analyse relationships between drifts in a consensus score such as CEM and in variable importance, followed by confirmation of any detected drifts using actual dataset trends.

## Limitations

Although statistical rigour has been applied to determine whether performance drift is a barrier to clinical risk modelling and decision-making, further work could be done to apply more statistically sensitive approaches to comparing the interactions of trends in dataset drift, performance drift and variable importance drift. While CEM is a consensus score that enhances clinical evaluation of complex relationships across different aspects of model performance, compressing the net benefit measure into a single value would mean that further decision curve analysis may be required if individual-specific threshold-based decisions were to be fully considered.

## CONCLUSION

This study addresses the gap in understanding the effect of dataset drift on the performance of ML and traditional models over time, which presents a barrier to the clinical application of ML. This was demonstrated by highlighting the importance of using a temporal and non-temporal ranking invariant consensus-based score for evaluating various ML approaches against traditional models, using smoothed and unsmoothed trend analysis, while comparatively assessing for relationships between and within variable importance drift, performance drift and actual dataset drift, for selecting the clinical ML model that minimizes the risk of patient harm. The strong evidence of all models showing a decrease in at least 3 of the 5 individual metrics within CEM demonstrates the importance of clinicians and ML governance teams actively monitoring the effects of dataset drift on models that are prepared for or being used for cardiac surgery risk prediction. Our data suggest that EuroSCORE II should be replaced with better performing ML models such as Xgboost and RF, which have demonstrated less drift over time. Future work will be required to determine the interplay between Xgboost and RF and whether ensemble models could take advantage of their respective performance advantages.

## Data Availability

All data used in this study are from the National Adult Cardiac Surgery Audit (NACSA) dataset. These data may be requested from Healthcare Quality Improvement Partnership (HQIP).

https://www.hqip.org.uk/national-programmes/accessing-ncapop-data/#.Ys6gN-zMLdp

## Abbreviations and Acronyms

AUC: area under receiver operating characteristic curve
CEM: Clinical Effective Metric
ECE: Expected Calibration Error
ES II: Euroscore II
AI: Artificial intelligence
ML: machine learning
RF: random forest
NN: Neural Network (Neuronetwork)
SVM: support vector machine
XGBoost: extreme gradient boosted trees Ensemble using several models to derive a consensus prediction
SHAP: (SHapley Additive exPlanations)

## Acknowledgement

This work was supported by a grant from the BHF-Turing Institute and the NIHR Biomedical Research Centre at University Hospitals Bristol and Weston NHS Foundation Trust and the University of Bristol.

## Data availability

All data used in this study are from the National Adult Cardiac Surgery Audit (NACSA) dataset. These data may be requested from Healthcare Quality Improvement Partnership (HQIP), https://www.hqip.org.uk/national-programmes/accessing-ncapop-data/#.Ys6gN-zMLdp.

